# Engagement in antenatal and HIV care among pregnant women before and after Option B+ policy implementation in South Africa

**DOI:** 10.1101/2024.10.31.24316487

**Authors:** Candice Hwang, Nelly Jinga, Mukesh Dheda, Otty Mhlongo, Pinky Phungula, Kate Clouse, Mark D. Huffman, Matthew P. Fox, Mhairi Maskew

**Affiliations:** Health Economics and Epidemiology Research Office, Faculty of Health Sciences, University of the Witwatersrand, Johannesburg, South Africa; Department of Internal Medicine, Stanford University School of Medicine, Stanford, California, USA; National Department of Health Pharmacovigilance Centre for Public Health Programmes, Pretoria, South Africa; Kwa Zulu Natal Department of Health, KwaZulu-Natal, South Africa; Vanderbilt University School of Nursing, Nashville, Tennessee, USA; Vanderbilt Institute for Global Health, Vanderbilt University Medical Center, Nashville, Tennessee, USA; Division of Cardiology, Department of Medicine, Washington University School of Medicine, St. Louis, Missouri, USA; The George Institute for Global Health, University of New South Wales, Sydney, Australia; Department of Global Health, Boston University School of Public Health, Boston, Massachusetts, USA; Department of Epidemiology, Boston University School of Public Health, Boston, Massachusetts, USA

**Author notes:** **Corresponding author:** Nelly Jinga.

**Keywords:** Antenatal, mother-to-child transmission, HIV, pregnancy, women, antiretroviral therapy, Option B

## Abstract

**Background:** Substantial gains have been made in South Africa in the prevention of vertical transmission of HIV over the past decade.

**Objectives:** to determine whether engagement in antenatal and HIV care among pregnant Women Living with HIV (WLWH) differed after Option B+ implementation.

**Methods:** We analysed cohort data from a pregnancy and birth defects surveillance system in KwaZulu-Natal (KZN). We report on two co-primary outcomes related to engagement in HIV care: 1) timing and number of ANC visits during the pregnancy period; and 2) timing of ART initiation (both self-reported ART use in interviews and observed initiation of treatment in maternal records). The association of policy era on the timing of ANC presentation was assessed using log-binomial regression modelling. We also report proportions initiating ART before or during pregnancy stratified by policy era.

**Results:** Data from 40,357 women, including 16,016 (40%) WLWH were analysed. During the Option B+ era, 24% of pregnant WLWH attended their first antenatal care visit during the first trimester, compared to 16% during the Option B era (RR=1.52;95%CI=1.41-1.64). The proportion of women living with HIV who initiated ART prior to pregnancy was also higher during the Option B+ era compared to the Option B era, though this result was limited by missingness in the data.

**Conclusions:** Engagement in antenatal and HIV care improved after Option B+ implementation. In the Option B+ era, South Africa has made significant progress towards the goal of eliminating mother-to-child transmission of HIV.

**What this study adds:**

- There has been an increase in the proportion of pregnant WLWH initiating ART prior to pregnancy and presenting for antenatal care (ANC) during the first trimester.
- The findings suggest improved timing and frequency of ANC visits during pregnancy, moving towards the UNAIDS 2025 targets.

## INTRODUCTION

Substantial gains have been made in South Africa in the prevention of vertical transmission of HIV over the past decade but elimination of maternal-to-child transmission of HIV (MTCT) has not yet been achieved. The national estimate of early mother-to-child transmission of HIV was 2.6% in 2012-2013.(1) By 2015, it was reduced to 1.4% at 6 weeks postpartum and 2% at 18 months.(2) National surveys by the South African Medical Research Council (SAMRC) and Centers for Disease Control and Prevention showed that between 2010 and 2015, the prevention of mother-to-child transmission of HIV (PMTCT) programme in South Africa prevented about 80 000 – 85 000 newborn infections per year (2).

South Africa’s policy for providing antiretroviral therapy (ART) to pregnant women living with HIV (WLWH) has evolved over time, following World Health Organization (WHO) recommendations (**Table 1**). In April 2008, a dual-therapy PMTCT programme was implemented through which women were offered zidovudine starting at 28 weeks’ gestation or ART at a CD4 count of < 200 cells/µL.(3) In 2010, Option A was introduced with zidovudine given starting at 14 weeks’ gestation for women with a CD4 count of >350 cells/µL or triple-drug ART for women with a CD4 count of ≤350 cells/µL.(4) Option A was replaced in 2013 by the WHO-recommended Option B approach, where women were offered triple-drug therapy, with postpartum withdrawal of antiretrovirals after cessation of breastfeeding for those ineligible for lifelong treatment.(5) The progression from Option B to Option B+ in 2015 where pregnant WLWH were offered lifetime ART regardless of CD4 count, was one of the key developments in the South African national HIV policy.(6)

**Table 1.** Definitions of Option A, B and B+ policies and years implemented in South Africa.(7)

| PMTCT<br>POLICY AND<br>YEAR<br>IMPLEMENTED | MOTHER RECEIVES: |  | INFANT RECEIVES: |
| --- | --- | --- | --- |
| | Treatment (CD4<br>count ≤350<br>cells/mm<sup>3</sup>) | Prophylaxis (CD4<br>count >350 cells/mm<sup>3</sup>) | |
| **Option A (2008)** | Triple ARVs starting as soon as diagnosed, continued for life | **Antepartum:** AZT starting as early as 14 weeks gestation<br>**Intrapartum:** at onset of labor, sdNVP and first dose of AZT/3TC<br>**Postpartum:** daily AZT/3TC through 7 days postpartum | Daily NVP from birth through 1 week beyond complete cessation of breastfeeding; or through age 4–6 weeks, if not breastfeeding or if mother is on treatment |
| **Option B (2013)** | Triple ARVs starting as soon as diagnosed, continued for life | Triple ARVs starting as early as 14 weeks gestation and continued intrapartum and through childbirth if not breastfeeding or until 1 week after cessation of all breastfeeding | Daily NVP or AZT from birth through age 4–6 weeks, regardless of infant feeding method |
| **Option B+ (2015)** | Same for treatment and prophylaxis:<br>Regardless of CD4 count, triple ARVs starting as soon as diagnosed, continued for life |  | Daily NVP or AZT from birth through age 4–6 weeks regardless of infant feeding method |

\*Abbreviations: ARV = antiretroviral drugs, AZT = Zidovudine, 3TC = Lamivudine, NVP = Nevirapine

An effective PMTCT programme ideally engages with WLWH as early as possible during pregnancy, because increased time on ART before delivery is associated with reduced risk of vertical transmission of HIV in-utero, during labor and delivery, and during breastfeeding.(8) WHO guidelines for high-quality antenatal care (ANC) in low- and middle-income countries specify that the first contact should occur within the first trimester and there should be a minimum of eight contacts. Prior to 2016, the WHO recommended minimum number of ANC contacts was four (9).

We assessed the association of implementation of the Option B+ policy on engagement in antenatal and HIV care among a cohort of pregnant WLWH in the KwaZulu-Natal (KZN) province of South Africa. Here, we compare the timing of antenatal care and ART initiation during the periods before and after Option B+ implementation to assess South Africa’s progress towards achieving the UNAIDS 2025 goals related to pregnant women, specifically having 90% of pregnant WLWH initiated on ART before their current pregnancy (10).

## METHODS

### Setting and population

The Understanding Birth Outcomes for Mothers and Infants (UBOMI) cohort was initially developed by the South African National Department of Health, in partnership with the KZN Provincial Department of Health, to assess the risk of congenital malformations and adverse birth outcomes from exposure to antiretroviral therapy (ART) during pregnancy.(11) The majority of data was collected at Prince Mshiyeni Memorial Hospital, a large public tertiary healthcare center in Durban providing care for approximately 14,000 deliveries each year.

We analyzed data from the UBOMI cohort of all pregnant women who presented at Prince Mshiyeni Memorial Hospital for delivery and at three obstetric units within the hospital’s catchment area for antenatal care from October 2013 to August 2017. The study period overlapped the transition from Option B to B+ implementation. Trained surveillance nurses interviewed pregnant women at their first antenatal care visit and obtained information on demographics, health and health-seeking behaviors during pregnancy, obstetric and neonatal history, HIV status and ART history, and maternal disease and substance exposures. These data were collected via a standardized digital form. Study enrolment was initially limited to weekdays due to limited data collector availability in the first year of the study, and in subsequent years study enrolment was expanded to every day. Consent was waived because these were routinely-collected data gathering for the national pregnancy exposure and birth defects surveillance system. Maternal case record data at the time of delivery were used to confirm and expand on what was reported in the interview. Gestational age at birth was routinely documented by nursing staff based on the estimated date of delivery calculated during the first ANC visit using last menstrual period (LMP), and confirmed by ultrasound, when available.

### Study Variables

We defined the Option B era as the period from October 2013 to December 2014 while the Option B+ era was defined as the period from January 2015 to August 2017. We report on two co-primary outcomes related to engagement in HIV care: 1) timing and number of ANC visits during the pregnancy period; and 2) timing of ART initiation (both self-reported ART use in interviews and observed initiation of treatment in maternal records).

### Statistical Analysis

The characteristics of women who engaged in HIV care and the timing and frequency of their participation in antenatal care, before and after the implementation of Option B+ are described using frequencies and simple proportions and are stratified by policy era. The association between policy era and timing of ANC presentation was assessed using log-binomial regression modelling. Estimated risk differences (RD), crude relative risks (RR) with 95% confidence intervals (CI) were reported. We also reported proportions initiating ART before or during pregnancy by policy era. Results are first reported with missing data excluded, which are then addressed with best and worst case sensitivity analysis in the discussion.

### Ethical Considerations

Approval for this surveillance system was obtained from the South African Medical Research Council’s ethics committee (protocol EC015-7/2013 approved on 30 July 2013). As this provincial surveillance system has since evolved to become a national sentinel surveillance system, consent for routine data gathering was not required, but digital photography of infants with congenital malformations, as well as stillbirths, which is not routine, required explicit consent. Analyses of de-identified data was also approved by the Human Research Ethics Committee of the University of the Witwatersrand (Medical) under protocol M200237 (approved 15 June 2020).

## RESULTS

A total of 40,318 HIV positive and HIV negative pregnant women with a median age of 24 years (IQR 20-30) attended ANC at Prince Mshiyeni Memorial Hospital from October 2013 to August 2017. Of these, 15,988 (39.7%) were WLWH, 17 of whom could not be assigned to a policy era because of missing data collection dates. Table 2 summarizes the characteristics of this study population stratified by PMTCT policy eras under study. There were no meaningful differences between groups in terms of education, employment status, age, gravidity and parity. However, a higher proportion of WLWH received ART during the Option B+ era (98%) compared to the period before Option B+ (92%).

**Table 2.** Demographics of WLWH who attended antenatal care at Prince Mshiyeni Memorial Hospital from October 2013 to August 2017 by policy era.

| Characteristic | Prior to Option B+<br>(October 2013 to<br>December 2014) (n =<br>5,483) | Option B+ (January 2015<br>to August 2017) (n =<br>10,498) |
| --- | --- | --- |
| **Education level, n (%)** |  |  |
| None | 14 (0.3%) | 10 (<0.1%) |
| Primary | 455 (8.3%) | 239 (2.3%) |
| Secondary | 4,781 (87%) | 9,811 (93%) |
| University/Tertiary | 209 (3.8%) | 350 (3.3%) |
| Missing | 24 (0.4%) | 88 (0.8%) |
| **Employment status, n (%)** |  |  |
| Employed | 932 (17%) | 1,715 (16%) |
| Unemployed | 4,527 (83%) | 8,727 (83%) |
| Missing | 24 (0.4%) | 56 (0.5%) |
| **Age, Median (IQR)** | 27 (23 – 32) | 28 (24 – 32) |
| <20 | 480 (8.8%) | 741 (7.1%) |
| 20-34 | 4,215 (77%) | 7,926 (76%) |
| ≥35 | 786 (14%) | 1,623 (15%) |
| Missing | 2 (<0.1%) | 208 (2.0%) |
| **Gravidity, Median (IQR)** | 2 (2 – 3) | 2 (2 – 3) |
| 1 | 1,181 (22%) | 2,044 (19%) |
| 2 | 2,233 (41%) | 3,972 (38%) |
| ≥3 | 2,068 (38%) | 4,347 (41%) |
| Missing | 1 (<0.1%) | 135 (1.3%) |
| **Parity, Median (IQR)** | 1 (1 – 2) | 1 (1 – 2) |
| 0 | 1,329 (24%) | 2,331 (22%) |
| 1 | 2,285 (42%) | 4,193 (40%) |
| ≥2 | 1,868 (34%) | 3,839 (37%) |
| Missing | 1 (<0.1%) | 135 (1.3%) |
| **Receiving ART, n (%)** |  |  |
| Yes | 5,031 (92%) | 10,268 (98%) |
| No | 79 (1.4%) | 86 (0.8%) |
| Not Applicable | 0 (0%) | 0 (0%) |
| Missing | 373 (6.8%) | 144 (1.4%) |

*\*Data for 17 women in this cohort could not be assigned to a policy era because of missing data collection dates*
*\*\*IQR – Interquartile range*

### Uptake and engagement in antenatal care across policy eras

Table 3 reports uptake and engagement in ANC across policy eras. Median gestational age at first ANC visit decreased by 2 weeks during the Option B+ era; for HIV-negative women median gestational age at entry to ANC decreased from 21 to 19 weeks while among HIV-positive pregnant women, gestational age at ANC entry decreased from 20 weeks under Option B to 18 weeks under Option B+. Over 60% of women during both eras, regardless of HIV status, presented for their first ANC visit in their second trimester. The median of five ANC visits during pregnancy was consistent across policy eras and HIV status.

**Table 3.**
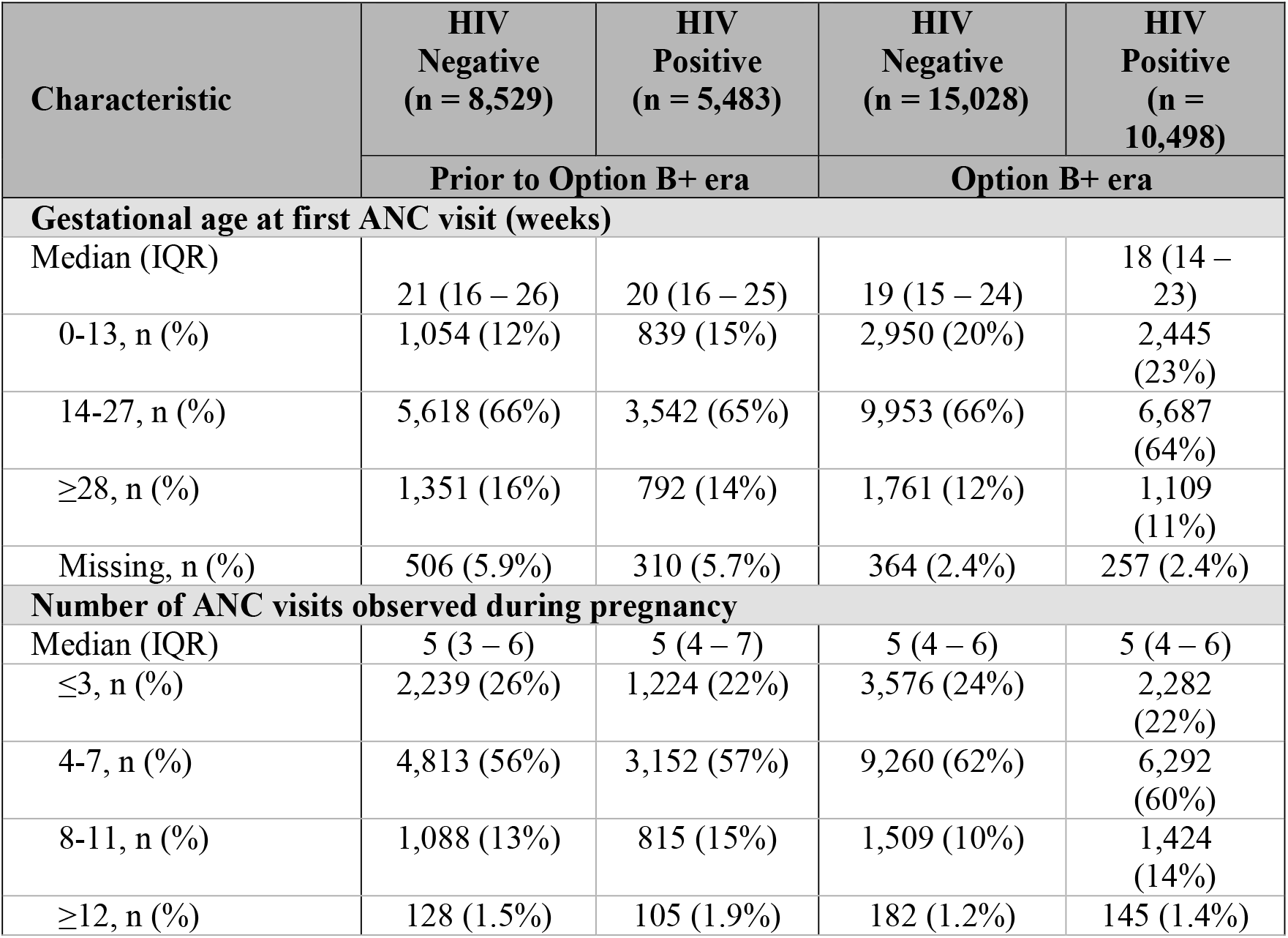
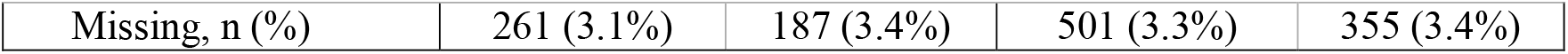
Uptake and engagement in antenatal care across policy eras.

| Characteristic | HIV<br>Negative<br>(n = 8,529) | HIV<br>Positive<br>(n = 5,483) | HIV<br>Negative<br>(n = 15,028) | HIV<br>Positive<br>(n = 10,498) |
| --- | --- | --- | --- | --- |
|  | Prior to Option B+ era |  | Option B+ era |  |
| Gestational age at first ANC visit (weeks) |  |  |  |  |
| Median (IQR) | 21 (16 – 26) | 20 (16 – 25) | 19 (15 – 24) | 18 (14 – 23) |
| 0-13, n (%) | 1,054 (12%) | 839 (15%) | 2,950 (20%) | 2,445 (23%) |
| 14-27, n (%) | 5,618 (66%) | 3,542 (65%) | 9,953 (66%) | 6,687 (64%) |
| ≥28, n (%) | 1,351 (16%) | 792 (14%) | 1,761 (12%) | 1,109 (11%) |
| Missing, n (%) | 506 (5.9%) | 310 (5.7%) | 364 (2.4%) | 257 (2.4%) |
| Number of ANC visits observed during pregnancy |  |  |  |  |
| Median (IQR) | 5 (3 – 6) | 5 (4 – 7) | 5 (4 – 6) | 5 (4 – 6) |
| ≤3, n (%) | 2,239 (26%) | 1,224 (22%) | 3,576 (24%) | 2,282 (22%) |
| 4-7, n (%) | 4,813 (56%) | 3,152 (57%) | 9,260 (62%) | 6,292 (60%) |
| 8-11, n (%) | 1,088 (13%) | 815 (15%) | 1,509 (10%) | 1,424 (14%) |
| ≥12, n (%) | 128 (1.5%) | 105 (1.9%) | 182 (1.2%) | 145 (1.4%) |
| Missing, n (%) | 261 (3.1%) | 187 (3.4%) | 501 (3.3%) | 355 (3.4%) |

### Early presentation for antenatal care

We also assessed the timing of ANC presentation, excluding the 17 women with unknown presentation dates (**Table 4**). During the Option B era, 13% of women living without HIV and 16% of WLWH presented for ANC in the first trimester, compared to 20% of women living without HIV and 24% of WLWH during the Option B+ era. Compared to women living without HIV in the Option B era, women living without HIV under Option B+ were 53% more likely to present for ANC in the first trimester (RR 1.53, 95% CI: 1.44-1.63). In both eras, WLWH were more likely to present for ANC in the first trimester compared to women living without HIV (Option B: RR 1.23, 95% CI: 1.14-1.34; Option B+: RR 1.82, 95% CI: 1.70-1.94).

**Table 4.** First Trimester Presentation to ANC, excluding women with unknown presentation date.

| Maternal HIV status and policy era | Overall (n) | Women presenting to ANC during first trimester ANC (n, %) | Risk Difference | Relative Risk (95% CI) |
| --- | --- | --- | --- | --- |
| **HIV-negative** |  |  |  |  |
| Option B | 8,023 | 1,054 (13%) | Ref | Ref |
| Option B+ | 14,664 | 2,950 (20%) | 0.07 | 1.53 (1.44-1.63) |
| **HIV-Positive** |  |  |  |  |
| Option B | 5,173 | 839 (16%) | 0.03 | 1.23 (1.14-1.34) |
| Option B+ | 10,241 | 2,445 (24%) | 0.11 | 1.82 (1.70-1.94) |

\* Totals exclude those with unknown presentation date

### Timing of ART initiation among pregnant women during the Option B and Option B+ eras

Across all study years, 4,603 (29%) of WLWH initiated ART before pregnancy, 7,525 (47%) initiated ART during pregnancy, 3579 (22%) initiated ART at an unknown date, 99 (1%) were never started on ART, and 175 (1%) were unknown. Figure 1 summarises timing of ART initiation in relation to pregnancy. The proportion initiating ART before or during pregnancy was high across both eras (>90% of those with known ART start dates), with 37% initiating prior to pregnancy among those with known ART start dates. Among women with known dates of initiation, 31% of WLWH initiated ART prior to pregnancy in the Option B era, compared to 53% WLWH in the Option B+ era.

**Figure 1.**
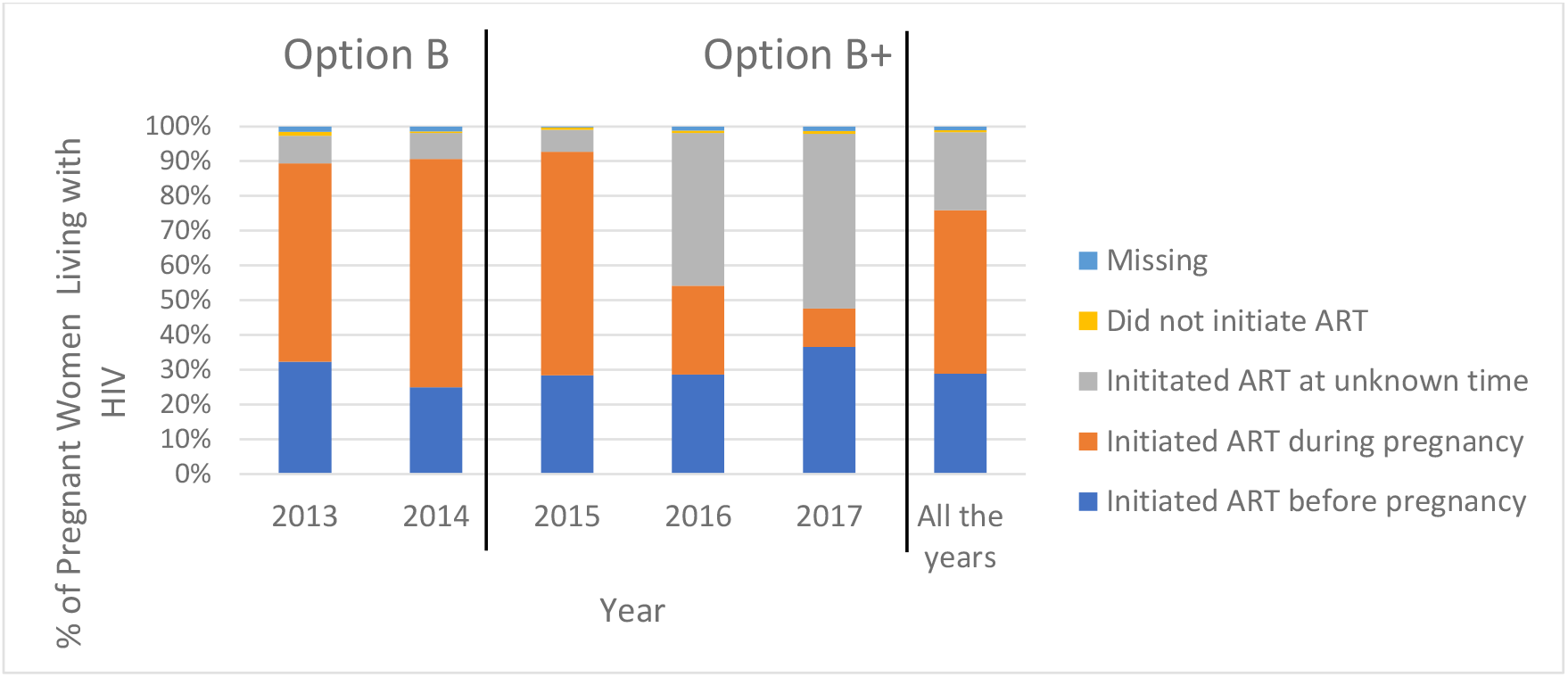
Timing of ART initiation relative to pregnancy for WLWH.

## DISCUSSION

As South Africa seeks to meet its vertical HIV transmission goals through policies of expanded access to ART, understanding the uptake and timing of engagement in HIV care remains critical. Here, we estimate engagement in antenatal care and uptake of HIV care before and after Option B+ implementation in South Africa between 2013-2017. We noted improvements in timing of entry to antenatal care. While overall less than one fifth presented for care during the recommended period in the first trimester, WLWH presented to ANC on average two weeks earlier in their pregnancy in the Option B+ era compared to the Option B era. In terms of frequency of care, almost 75% of women had four or more ANC visits, which is the minimum number of contacts during pregnancy recommended by the WHO prior to 2016.(9) We note, however, that these improvements in timing of entry to antenatal care also occurred among HIV negative women, though overall HIV-positive women tended to present at earlier gestational age. It is thus likely that factors other than implementation of the Option B+ policy have positively impacted the timing of entry to ANC.

Despite this caution, our findings are in keeping with other national estimates from the South African Demographic Health Survey (DHS).(12) DHS results aggregated over 1998 to 2016 noted timing of ANC presentation to be 17% in the first trimester, 72% in the second trimester, and 10% in the third trimester, similar to our findings. The DHS results also indicated 79% of women nationally and 83% of women in KZN attended at least four ANC visits during pregnancy, similar to the 76% of women in our study(13).

Most WLWH in this cohort were started on ART prior to delivery. We found that the proportion of WLWH with known ART initiation dates who initiated ART prior to their pregnancy was higher in the Option B+ era than the Option B era. Specifically, in 2017, excluding those with missing data, around 75% of WLWH in this cohort were on ART before their current pregnancy. In our results, 98% of WLWH across 2013-2017 received ART at some point before or during their pregnancy. This result is similar to what was found in the National Antenatal Sentinel HIV Survey, where 87% of women in 2017 knew their HIV-positive status, and 96% in 2019, were receiving ART at the time they were surveyed during pregnancy (14). These estimates are still short of the UNAIDS 2025 goal to have 90% of pregnant WLWH already be initiated on ART before conception (10).

Our findings should be considered in the context of some limitations with this analysis. First, the large proportion of missing ART initiation dates during the 2016-17 period limits our interpretation of these results. In the best case scenario (we assume all those with unknown ART initiation dates initiated ART before pregnancy) about half of the women initiated ART before pregnancy. In the worst case scenario (we assume all those with unknown ART initiation dates initiated during pregnancy) less than one third were already on ART at entry to ANC across all study years. Second, though a large dataset, women observed were from one catchment area of Prince Mshiyeni Hospital in KZN province, which may limit the generalizability of our findings beyond this population. Also, no viral load data was available in this analysis and thus, we were unable to assess whether the impact of Option B+ implementation extended beyond engagement in HIV care to sustained viral load suppression. Finally, for many women, breastfeeding continues for months after delivery and so those on Option B may still have been breastfeeding (and thus still on ART) by the time B+ was implemented in 2015. In this way, the distinction between the policy eras may not have been very clear among breastfeeding women and to the extent which this occurred, benefits of the implementation of Option B+ in our estimates would be under-estimated.

Despite these limitations, our findings from this large cohort offer valuable evidence of gains made in terms of meeting the UNAIDS 2025 targets. We found that South Africa has made progress in terms of improving women’s engagement in ANC and HIV care. Despite increases in prior ART use at entry to ANC, pregnancy services remain an important portal for access to HIV care. For those women accessing care, most receive antenatal service at the appropriate frequency, though late presentation for antenatal care persists beyond the recommended gestational age. As a result of this, critical gaps to achieving targets for engagement in HIV care among pregnant women remain.

## Author contributions

CH and MM conceptualized the study and methodology. CH and NJ performed the analysis. CH and NJ wrote the original draft. CH, NJ, MM, MF, KC, CN, MD, MH, PP, OM provided interpretation of results and critical review of the manuscript.

## Data availability statements

The data that support the findings of this study are owned by the KwaZulu-Natal Provincial Department of Health, and access is governed by policies and procedures in response to requests made directly to the Department. As such, the study teams will not have authority to release the data to the public or other data-sharing repositories. However, these data can be requested by the public through standardized request forms, which are then considered in an internal review procedure.

## Disclaimer

The views and opinions expressed in this article are those of the authors and are the product of professional research. It does not necessarily reflect the official policy or position of any affiliated institution, funder, agency, or that of the publisher. The authors are responsible for this article’s results, findings, and content.

